# Interventions Promoting Recovery from Depression for Patients Transitioning from Outpatient Mental Health Services to Primary Care: Protocol for a Scoping Review

**DOI:** 10.1101/2022.10.06.22280499

**Authors:** Anne Sofie Aggestrup, Frederik Martiny, Maria Faurholt-Jepsen, Morten Hvenegaard, Robin Christensen, Annette Sofie Davidsen, Klaus Martiny

**Affiliations:** Copenhagen Affective Disorder Research Centre (CADIC), New Interventions in Depression (NID) Group, Mental Health Centre Copenhagen, Department of Clinical Medicine, University of Copenhagen, Copenhagen, Denmark; The Research Unit for and Section of General Practice, Department of Public Health, University of Copenhagen, Copenhagen, Denmark; Center for Social Medicine, Bispebjerg and Frederiksberg Hospital, Copenhagen, Denmark; Copenhagen Affective Disorder Research Centre (CADIC), Mental Health Centre Copenhagen, Copenhagen University Hospital, Rigshospitalet, Copenhagen, Denmark; Competence Centre for Rehabilitation and Recovery, Mental Health Centre Ballerup, Ballerup, Denmark; Section for Biostatistics and Evidence-Based Research, the Parker Institute, Bispebjerg and Frederiksberg Hospital & Department of Clinical Research, University of Southern Denmark, Odense University Hospital, Denmark

**Keywords:** Depression, recovery, transitioning, mental health services, primary care

## Abstract

**Introduction:** Patients with severe Major Depressive Disorder (MDD) have an increasing risk of new psychiatric hospitalizations following each new episode of depression highlighting the recurrent nature of the disorder. Furthermore, patients are not fully recovered at the end of their treatment in outpatient mental health services, and residual symptoms of depression might explain why patients with MDD have a high risk of relapse. However, evidence of methods to promote recovery after discharge from outpatient mental health services is lacking. The proposed scoping review aims to systematically scope, map and identify the literature and knowledge gaps on existing interventions that promote recovery from MDD for patients transitioning from outpatient mental health services to primary care.

**Methods and analysis:** The proposed scoping review will follow the latest methodological guidance by the Joanna Briggs Institute (JBI) in tandem with the Preferred Reporting Items for Systematic reviews and Meta-Analysis - extension for Scoping Reviews (PRISMA-ScR) checklist. The review is ongoing. Four electronic databases (Medline via PubMed, PsycINFO, CINAHL, and Sociological Abstracts) were systematically searched from 20 January 2022 till 29 March 2022 using keywords and text words. The review team consists of three independent screeners. Two screeners have completed the initial title and abstract screening for all studies retrieved by the search strategy. Currently, we are in the full text screening phase. Reference lists of included studies will be screened, and data will be independently extracted by the review team. Results will be analyzed qualitatively and quantitatively.

**Ethics and dissemination:** The chosen methodology is based on the use of publicly available information and does not require ethical approval. Results will be published in an international peer reviewed scientific journal and additionally shared with relevant local and national authorities.

**Registration:** Following publication, we intend to register the protocol on Open Science Framework.

**Data availability statement:** Data sharing not applicable as no datasets generated and/or analyzed for this study.

**Strengths and limitations of this study:** To our knowledge, this scoping review is the first to identify and map interventions that aim to promote recovery from severe major depressive disorder for patients transitioning from outpatient mental health services to primary care.

The proposed scoping review will be conducted in accordance with the Preferred Reporting Items for Systematic reviews and Meta-Analysis - extension for Scoping Reviews in tandem with the latest framework for scoping review proposed by the Joanna Briggs Institute.

The review will not assess the quality of intern validity of included studies. However, we will outline the key characteristics of the best-available evidence in the area and comment of the applicability of the evidence in various settings.

## INTRODUCTION

### Disease Burden of Major Depressive Disorder and Treatment across Sectors

Major Depressive Disorder (MDD) is one of the most prevalent mental disorders worldwide with a lifetime risk of 20% for adults on a global level [1-3]. Approximately 5% of the general population experiences a depressive episode within a 12-month period [1-3]. Current predictions by the World Health Organization indicate that by 2030 depression will be the leading cause of disease burden globally [4]. MDD has a negative impact on quality of life, and reduces psychosocial, social, and occupational functioning, and markedly increases morbidity and mortality [5-7].

A huge body of evidence from e.g., epidemiological surveys has documented a strong interconnection and increased comorbidity of MDD with other mental disorders, most notably with anxiety disorders and substance use disorders [5, 8-11]. MDD has furthermore been associated with comorbid physical diseases, e.g., diabetes and heart diseases [5, 6, 12-15] and social difficulties [10, 16, 17], e.g., poor work participation, drift to a lower social class, and poorer education.

Thus, MDD is a major burden for the individual patient and public health throughout the world [1, 2, 18, 19], and on a societal level, MDD leads to significant direct costs for treatment, care and rehabilitation and indirect costs due to disease-related work disability and mortality [5, 6, 12-15].

A diagnosis of MDD is reached when patients experience five or more out of nine symptoms during the same 2-week period and at least one of these symptoms should be either depressed mood or loss of interest and pleasure [20]. In addition, patients with MDD experience a variety of associated emotional, cognitive, and behavioral symptoms [10, 21, 22].

Most patients with MDD are diagnosed and treated in primary care by general practitioners, but research has shown that the ability to detect, diagnose, and treat patients with MDD is often insufficient [23-25]. Furthermore, there is sparse evidence to conclude which type of treatment approach is most effective in preventing relapse or recurrence of MDD [26, 27]. A Cochrane review concluded that patients taking antidepressant medication were less likely to relapse or to experience a recurrent episode compared to patients not taking antidepressant medication (13.9% versus 33.8%) [27]. There are, however, methodological problems in assessing the prophylactic effect of antidepressants [28]. There is also some evidence that non-pharmacological treatment options can induce a reduction in depression symptoms and further remission, i.e., rumination-group cognitive-behavioral therapy, light, exercise, and sleep regulation therapy [29, 30].

MDD is often reoccurring and in some cases it becomes chronic. After treatment of the first episode of severe MDD, more than 50% of all inpatients will relapse [15, 31-36]. These residual symptoms of MDD and incomplete recovery are thus considered a significant contributor to the high risk of relapse for patients with MDD [37]. Therefore, MDD requires long-term and adequate multimodal treatment to induce recovery and prevent/reduce the risk of further episodes. Specialized mental health services typically manage treatment of severe recurrent depression and difficult to treat depression or pressing suicidal ideation [38-40]. However, most mental health services only offer treatment for shorter periods, and research concludes that too early discharge can remove critical support and treatment from vulnerable patients that are not fully recovered at the end of their treatment in outpatient mental health services [36, 41, 42]. In addition, research has shown that patients may not have full confidence in the general practitioners’ ability to decide continuation/discontinuation of antidepressant due to a perceived lack of knowledge and time in general practice [38, 43]. As described, some patients relapse in the treatment gaps between outpatient mental health services and often insufficient treatment in primary care. Patients with MDD require ongoing maintenance treatments over the long term to facilitate continued recovery.

### Recovering from Major Depressive Disorder

The concept of recovery was first used in the 1960s, primarily aimed to restore human rights as part of user movements responding to the perceived dominant, and stigmatizing notion of mental illness as chronic with little possibility for improvement [44]. Since then, recovery has become an increasingly important aspect of mental healthcare [45, 46]. The main notions of recovery in mental health literature are the concepts of *clinical* - and *personal* recovery.

Clinical recovery refers to a process of individual recovery from mental illness by remission from symptoms and attainment of functional improvement [47-49]. Personal recovery refers to a process in which the individual recovers from the social consequences of the mental illness, thus regaining a meaningful life and participating in the community by overcoming the challenges of mental illness with or without symptoms. Personal recovery is commonly conceptualized via the ‘CHIME’ Framework. It consists of five interrelated processes: **C**onnectedness with other people and the community; **H**ope and optimism about the future; overcoming stigma and redefining a positive sense of **I**dentity; **M**eaning in life as defined by rebuilding a meaningful life with social goals; **E**mpowerment, which includes taking personal responsibility and control over one’s life [50-57].

The two concepts of recovery have led to some polarization in the understanding of what recovery entails [58]. Recently, it has been argued that the two concepts should be considered complementary rather than contrasting, especially to prevent that patients are left in limbo in the, sometimes, polarized discussion between researchers and clinicians [47]. Professionals working in psychiatry tend to focus on clinical recovery [47-49, 58], while general practitioners tend to focus more on personal recovery, which aligns well with a generalist and person-centered rather than disease-centered approach to care [43, 52, 58]. Yet, patients want both clinical and personal recovery [58]. Therefore, in this review, we focus on both clinical- and personal recovery, collectively referred to as “recovery”.

### Existing Evidence on Recovery from Major Depressive Disorder for Patients Transitioning from Outpatient Mental Health Services to Primary Care

To our knowledge, there are no published scoping reviews that summarize the evidence for interventions aiming to promote continued recovery from MDD for patients transitioning from outpatient mental health services to primary care. Most studies investigating recovery interventions and / or relapse prevention from MDD have been undertaken in primary care [38, 59, 60]. Two reviews have a specific focus on developing recovery interventions, e.g., scoping the evidence for internet-based recovery-oriented interventions [61], or developing a proposed logic model, i.e., a visual representation of what works for whom, why, and under which circumstances, for how recovery-oriented interventions could contribute to recovery [62]. However, these reviews were not specific to our target group of interest, did not focus on clinical recovery or patients transitioning from outpatient mental health services to primary care [61, 62]. Currently, the best available evidence in the field is a rapid review from 2021 by Blasi et al. [63] that identified practices for transitioning stable patients from outpatient mental health services to primary care, and a systematic review from 2006 by Gunn et al. [64] that assessed the effects of chronic illness management approaches for patients with depression in primary care. The rapid review by Blasi et al. [63] included 11 articles representing six categories of transition practices, with patient engagement as the most commonly described transition practice, followed by shared treatment planning, assessment of recovery and stability, care coordination, follow-up and support, and medication management. However, the review did not draw conclusions about best practices or the importance of specific transition processes or strategies, including interventions that promotes recovery for patients transitioning. In addition, the authors may have missed some relevant articles due to the rapid review timeline for literature searching and study selection [63]. The systematic review by Gunn et al. [64] found that system level interventions in primary care can led to a modest increase in recovery from depression. Yet, the quality of the evidence was poor and ten of the 11 randomized controlled trials included in the review (91%) were from the United States of America [64]. Thus, the authors concluded that possibly the findings in the reviews were likely not applicable to countries with strong primary care systems. Of note, the scope of the review was not recovery after discharge from mental health services. In addition, neither observational nor qualitative studies were included for review, limiting the review’s ability to provide a comprehensive overview of the field of recovery from MDD [64]. Lastly, much research on recovery from MDD has been conducted since 2006, making an updated review relevant [65, 66]. Nevertheless, it is plausible that shared care models for treatment of MDD between outpatient mental health services and primary care may improve recovery [67]. Therefore, we believe that a scoping review on this field will be valuable to identify knowledge gaps due to its connection with and to inform an ongoing co-design development project that we will describe briefly below.

### The Scoping Review Informs a Co-design Process that aims to Develop a Complex Intervention

Given high rates of relapse and residual symptoms for patients with MDD following transitioning from outpatient mental health services to primary care, new strategies to promote continued recovery are required. A promising method is to develop an intervention that promotes continued recovery from MDD for patients transitioning. This scoping review is one part of an ongoing stakeholder co-design project (supplementary file A) located in the Capital Region of Denmark. In the present review, stakeholders are involved in the design and conduct of the review. Other activities involved in the co-design process includes individual interviews, focus groups and workshops with stakeholders. The overall aim of the co-design project is to develop a complex intervention that promotes recovery from MDD for patients transitioning (supplementary file A) from outpatient mental health services to primary care. Following the development of the intervention, we plan to test the intervention over a series of feasibility studies [68-70].

### Objective

The proposed scoping review aims to systematically scope, map and identify the literature and knowledge gaps on existing interventions that promote recovery from MDD for patients transitioning from outpatient mental health services to primary care.

#### Research questions

*What characterizes interventions aiming to promote recovery from MDD for patients transitioning from outpatient mental health services to primary care, and under which circumstances do they work?*

## METHODS AND ANALYSIS

Scoping reviews are methodologically rigorous in their approach to examining the extent, range, and nature of research activity in a particular field. The methodology is particularly useful for identifying and synthesizing the best available evidence that spans a vast conceptual and methodological range in the health disciplines [71-75], as is the case within this research area. The first framework for conducting a scoping review was proposed by Arksey and O’Malley [71]. Extensions of this framework were later provided by Levac et al. [72]. These initial attempts have provided guidance to many researchers, but lack of methodological clarity continues to exist. In response to ongoing concerns about the scoping review methodology, the Joanna Briggs Institute (JBI) guidance for scoping reviews was developed by a working group of methodological experts and first published in 2015 [73]. JBI updated their guidance in 2017 and again in 2020 [74, 76].

This proposed scoping review will follow the latest methodological guidance by the JBI [77] in tandem with the Preferred Reporting Items for Systematic reviews and Meta-Analysis - extension for Scoping Reviews (PRISMA-ScR) checklist [78] (supplementary file B).

### Patient and public involvement

During protocol development, we used the TRANSFER approach [79] to involve relevant stakeholders in discussions about the scoping reviews’ aim and methods, aiming to promote relevance and transferability of the reviews’ findings. We included a diverse set of stakeholders over a series of meetings to gain perspectives from researchers from a) general practice, b) mental health services, and c) social medicine. We also conducted interviews with patients with MDD and a focus group with job consultants from the Municipality of Copenhagen to include their perceptions.

### Protocol and registration

This scoping review protocol is novel, i.e., not based on updates from previous review(s). A preprint will be registered at the medRxiv preprint server for health sciences.

Following publication, we intend to share the protocol and any supplementary material on Open Science Framework (OSF) (available at: https://osf.io/rr/). In case the conduct outlined in this protocol changes substantially during the review process, we will update the protocol in the OSF accordingly and report deviations from the protocol in the final publication(s).

### Eligibility criteria

The following eligibility criteria (table 1) guide the decision to in- or exclude studies identified for review. These are structured according to the PICOS acronym (population, intervention, comparator, outcome, and setting).

**Table 1.**
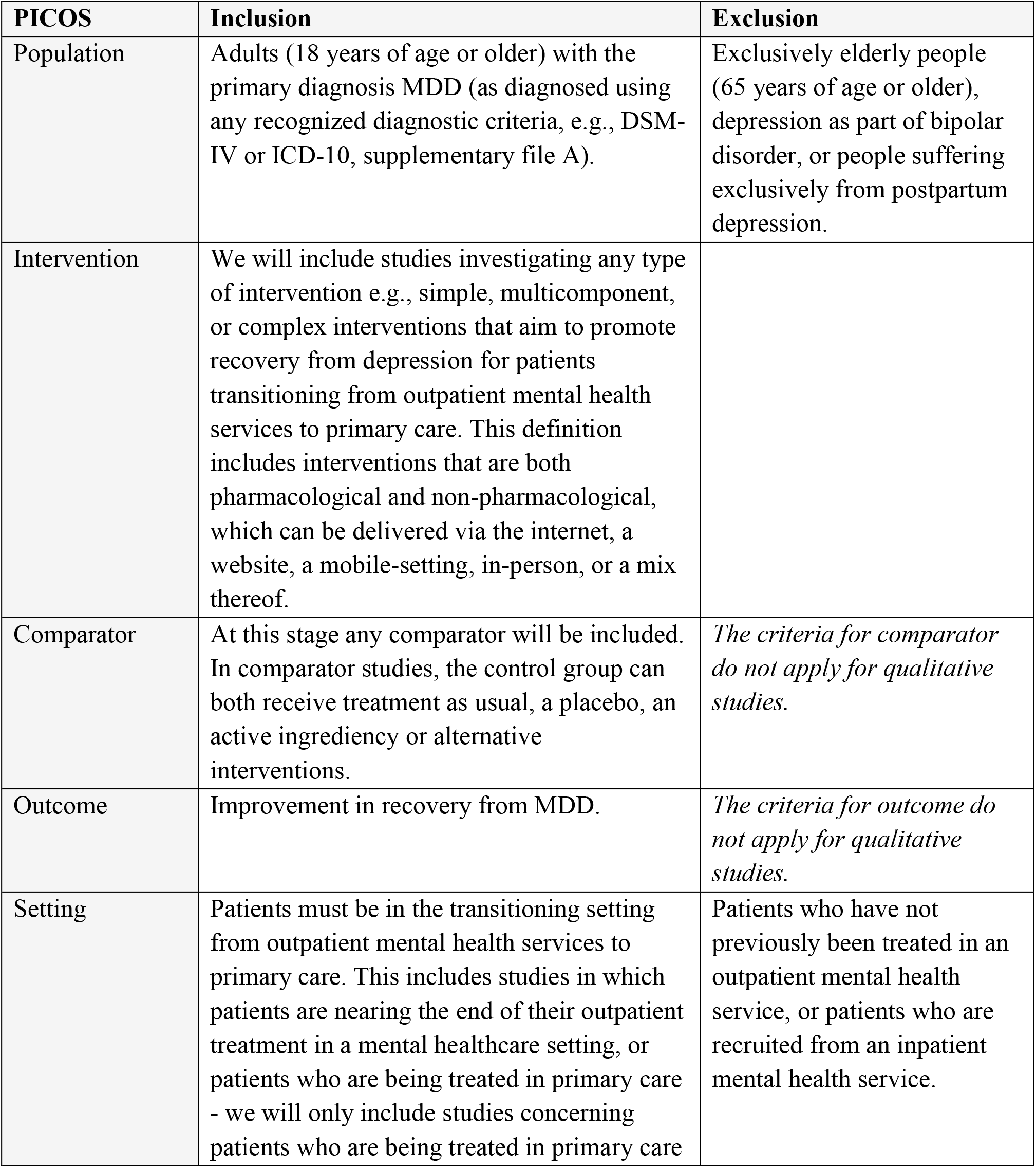
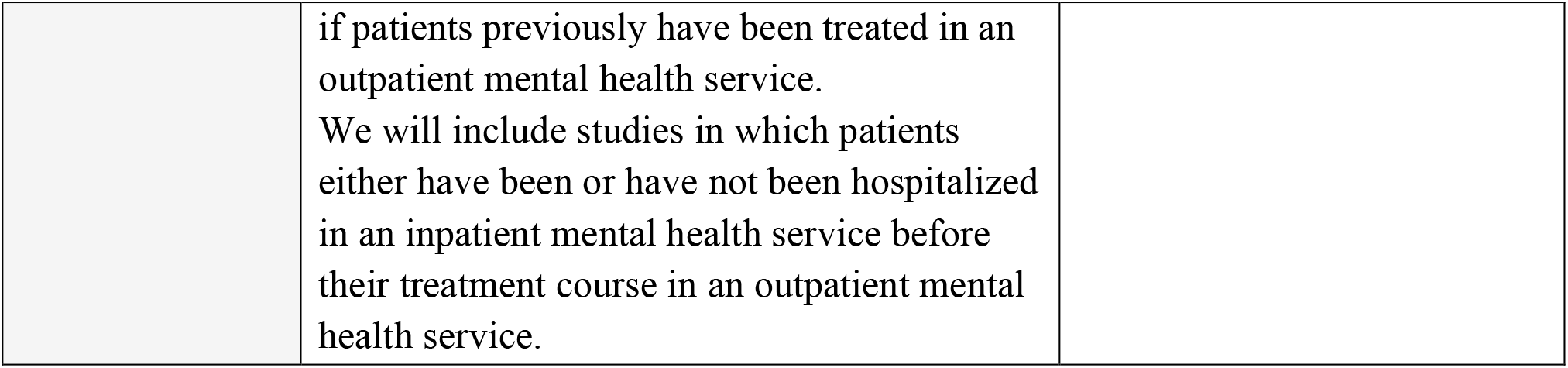
Eligibility criteria

| PICOS | Inclusion | Exclusion |
| --- | --- | --- |
| Population | Adults (18 years of age or older) with the primary diagnosis MDD (as diagnosed using any recognized diagnostic criteria, e.g., DSM-IV or ICD-10, supplementary file A). | Exclusively elderly people (65 years of age or older), depression as part of bipolar disorder, or people suffering exclusively from postpartum depression. |
| Intervention | We will include studies investigating any type of intervention e.g., simple, multicomponent, or complex interventions that aim to promote recovery from depression for patients transitioning from outpatient mental health services to primary care. This definition includes interventions that are both pharmacological and non-pharmacological, which can be delivered via the internet, a website, a mobile-setting, in-person, or a mix thereof. |  |
| Comparator | At this stage any comparator will be included. In comparator studies, the control group can both receive treatment as usual, a placebo, an active ingrediency or alternative interventions. | *The criteria for comparator do not apply for qualitative studies.* |
| Outcome | Improvement in recovery from MDD. | *The criteria for outcome do not apply for qualitative studies.* |
| Setting | Patients must be in the transitioning setting from outpatient mental health services to primary care. This includes studies in which patients are nearing the end of their outpatient treatment in a mental healthcare setting, or patients who are being treated in primary care - we will only include studies concerning patients who are being treated in primary care | Patients who have not previously been treated in an outpatient mental health service, or patients who are recruited from an inpatient mental health service. |
|  | if patients previously have been treated in an outpatient mental health service.<br>We will include studies in which patients either have been or have not been hospitalized in an inpatient mental health service before their treatment course in an outpatient mental health service. |  |

### Additional limits

No limits on publication date, language, country, or gender, no restrictions in type of study design. Both qualitative, quantitative and mixed-methods studies are included.

Articles without full text available will not be included.

### Information sources and search

The literature search was developed in collaboration with an information specialist with feedback from the stakeholders that were included via the TRANSFER [79] approach in discussions regarding the eligibility criteria (PICOS elements) for the review outlined above. We have searched the electronic databases of Medline via PubMed, PsycINFO, CINAHL, and Sociological Abstracts. The search strategy included both text words and Medical Subject Headings (MeSh)/Thesaurus headings terms. Prior to performing the search strategy, we searched for ongoing or completed scoping or systematic reviews in the area on Cochrane Library, Google Scholar, and the PROSPERO register to make sure there was not already relevant reviews in the area.

The search strategy for PubMed is available in supplementary file C. All databases were searched from 20 January 2022 to 29 March 2022.

Reference lists of included studies will be examined, i.e., backward citation tracing, to identify relevant studies potentially missed by the search strategy. Vice versa, we will do forward citation tracing of all included studies via Web of Science. The database searches will be re-run just before the final analysis is conducted to include the most recent evidence.

### Selection of sources of evidence

Results from the literature were exported from databases to the Covidence (https://www.covidence.org/) reference management software system. Duplication of database search results was removed used using EndNote 20 reference management software. Prior to the start of the review, all screeners were trained to use the Covidence system and received education about the content area, i.e., depression and recovery.

Relevant studies were screened through a two-step process for examining titles and abstracts and then full texts. The review team consists of three independent screeners. Two independent screeners completed the initial title and abstract screening for all studies retrieved by the search strategy. A third screener reviewed conflicts and resolved disagreements through discussion with the two other screeners. Over the course of two months, these unblinded screeners (unable to see each other votes in Covidence until they have cast their own, and vice versa, and they will not be blinded to the authors and journals) have screened 4605 titles and abstracts independently. Three screeners reviewed at least 1600 titles and abstracts each. Currently, we are in step two, the full text screening. Here, two independent screeners will review the full text of potentially eligible articles. Disagreements between screeners during full text screening will be resolved by discussion or, if needed, by consulting a third screener. If there is more than one article from the same study, the most updated data will be extracted. If information is missing or clarification of data is required, authors will be contacted via e-mail.

Overall reasons for inclusion/exclusion of studies will be documented and reported in a PRISMA flowchart [80] in the final article reporting the findings from the review process.

### Data charting process

The preliminary charting table (table 2) guides data extraction (charting). Design of the table was guided by/-inspired by the newest JBI guideline [77] and further developed for this scoping review in line with the review’s objectives and research questions in collaboration with stakeholders included via the TRANSFER approach [79]. Two screeners will independently extract data from included studies into a Microsoft Excel sheet organized in columns corresponding to the items in the table. The screeners will agree on revisions to the charting table as needed in an iterative process [81]. To ensure clarity and consistency between the screeners’ data extraction, and prior to initiating the full text article selection process, we will pilot test the data extraction process on a subset of potentially eligible full text articles. Screeners will resolve disagreements by discussion, and a third screener will adjudicate unresolved disagreements.

**Table 2.**
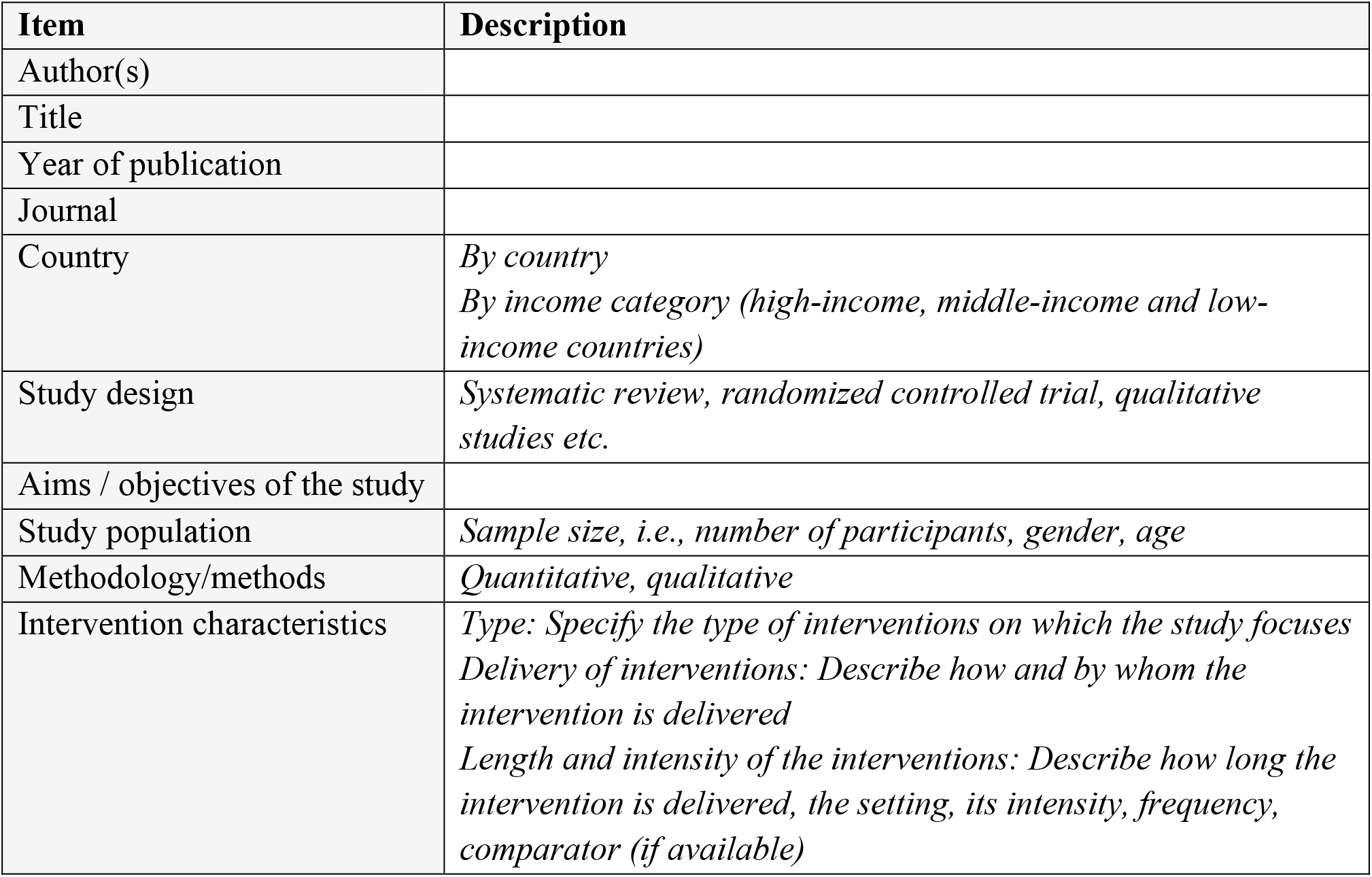
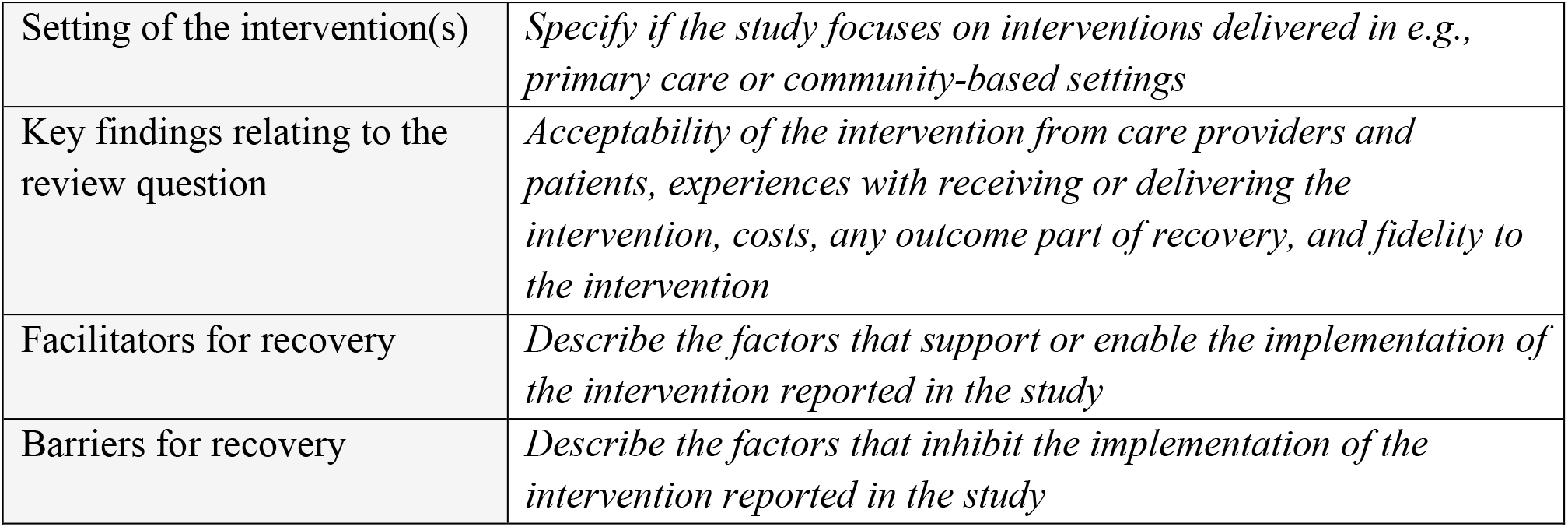
Preliminary charting table

| Item | Description |
| --- | --- |
| Author(s) |  |
| Title |  |
| Year of publication |  |
| Journal |  |
| Country | *By country*<br>*By income category (high-income, middle-income and low-income countries)* |
| Study design | *Systematic review, randomized controlled trial, qualitative studies etc.* |
| Aims / objectives of the study |  |
| Study population | *Sample size, i.e., number of participants, gender, age* |
| Methodology/methods | *Quantitative, qualitative* |
| Intervention characteristics | *Type: Specify the type of interventions on which the study focuses*<br>*Delivery of interventions: Describe how and by whom the intervention is delivered*<br>*Length and intensity of the interventions: Describe how long the intervention is delivered, the setting, its intensity, frequency, comparator (if available)* |
| Setting of the intervention(s) | *Specify if the study focuses on interventions delivered in e.g., primary care or community-based settings* |
| Key findings relating to the review question | *Acceptability of the intervention from care providers and patients, experiences with receiving or delivering the intervention, costs, any outcome part of recovery, and fidelity to the intervention* |
| Facilitators for recovery | *Describe the factors that support or enable the implementation of the intervention reported in the study* |
| Barriers for recovery | *Describe the factors that inhibit the implementation of the intervention reported in the study* |

### Data items

We will extract data as shown in table 2.

### Critical appraisal of individual sources of evidence

Since this is a scoping review, we will not conduct quality appraisal, which is consistent with the framework proposed by the JBI methodology for scoping reviews [74, 76].

### Synthesis of results

According to the JBI methodology [74, 76] for scoping reviews, the quantitative results extracted from included studies will be analyzed with descriptive statistics with visual representations of the data where possible, e.g., mapping the extracted data in a diagrammatic, tabular, or descriptive format. Qualitative findings from studies will be analyzed from a thematic perspective and, depending on the results, described regarding for example active ingredients, patient satisfaction, and barriers and facilitators for implementation.

The results will be classified under main conceptual categories, such as: “intervention type”, “duration of intervention”, “facilitators/barriers”, “aims”, “methodology adopted”, “key findings” (evidence established), and “gaps in the research field”. For each category reported, a clear explanation will be provided.

## Data Availability

Data sharing not applicable to this article as no datasets were generated or analysed during the current study.

## ETHICS AND DISSEMINATION

This scoping review constitutes the first step of a larger research project aiming to develop a complex intervention to promote recovery from MDD by optimizing the process of patients transitioning from outpatient mental health services to primary care. The chosen methodology is based on the use of publicly available information and does not require ethical approval. Results will be published in a peer-reviewed international scientific journal and shared with relevant local and national authorities via other communication channels.

## AUTHORS’ CONTRIBUTIONS

ASA drafted the protocol manuscript. All authors contributed to the development of the selection criteria, study selection, and screening strategy. ASA developed the initial search strategy with assistance of a science clinical librarian with expertise in review searching, and with feedback from the review authors. All authors read, provided feedback, and approved the final manuscript.

## SUPPLEMENTARY FILE A

## List of abbreviations / concepts

Abbreviation / concept: Definition
MDD: Major Depressive Disorder
DSM-IV: The Diagnostic and Statistical Manual of Mental Disorders, Fifth Edition
ICD-10: International Classification of Diseases and Related Health Problems 10^th^ Revision
Co-design: A participatory approach to design interventions in collaboration with stakeholders.
Stakeholder: Any individual or group who is responsible for or affected by health- and healthcare-related decisions that can be informed by research evidence [82]. In this study stakeholders are patients, general practitioners, psychiatrists, nurses, job-consultants, medical social workers, and researchers.
Patients transitioning: When patients move between care settings. In this study, we focus on patients’ transition from outpatient mental health services to primary care.

## SUPPLEMENTARY FILE B

**Preferred Reporting Items for Systematic reviews and Meta-Analyses extension for Scoping Reviews (PRISMA-ScR) Checklist**

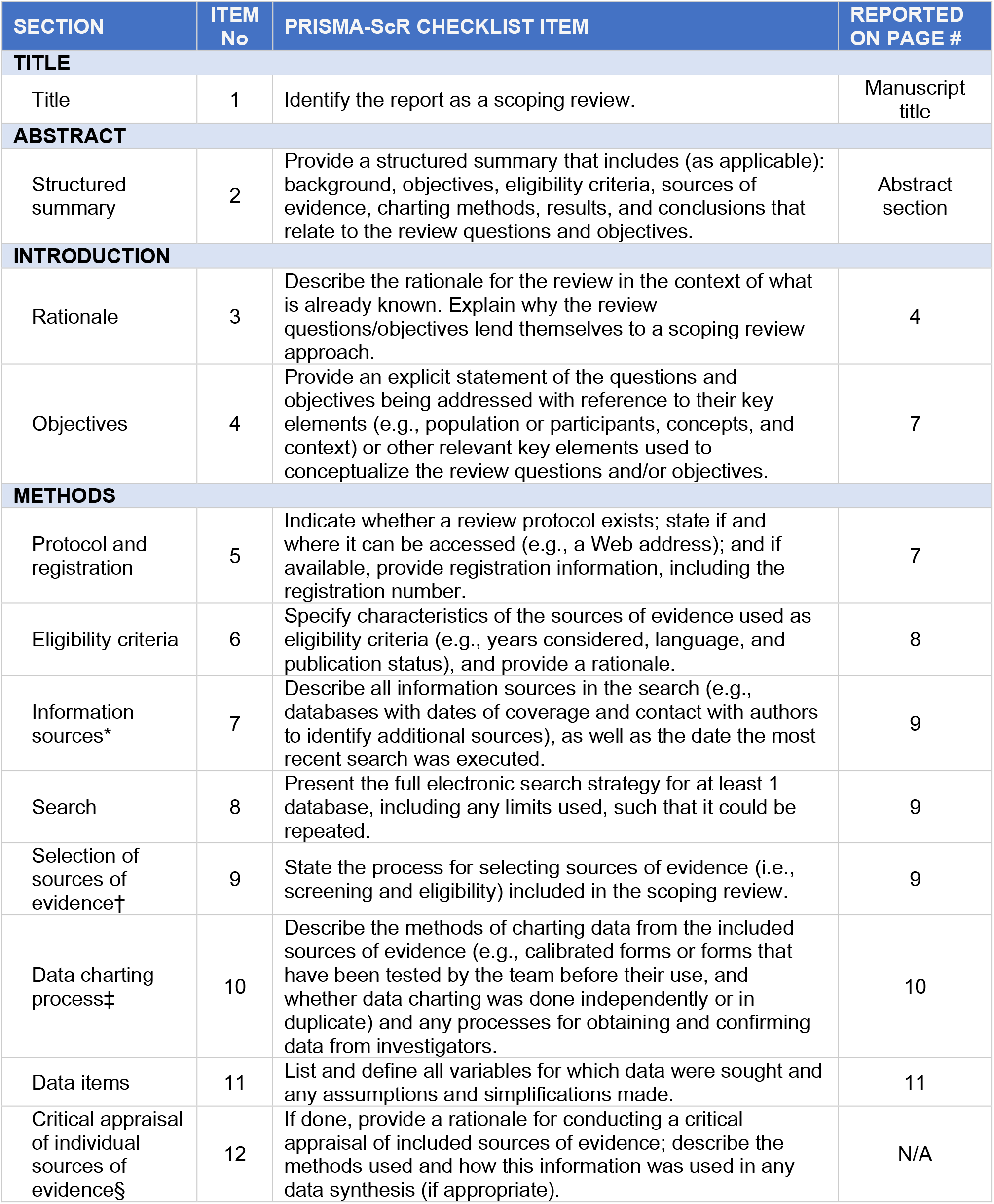
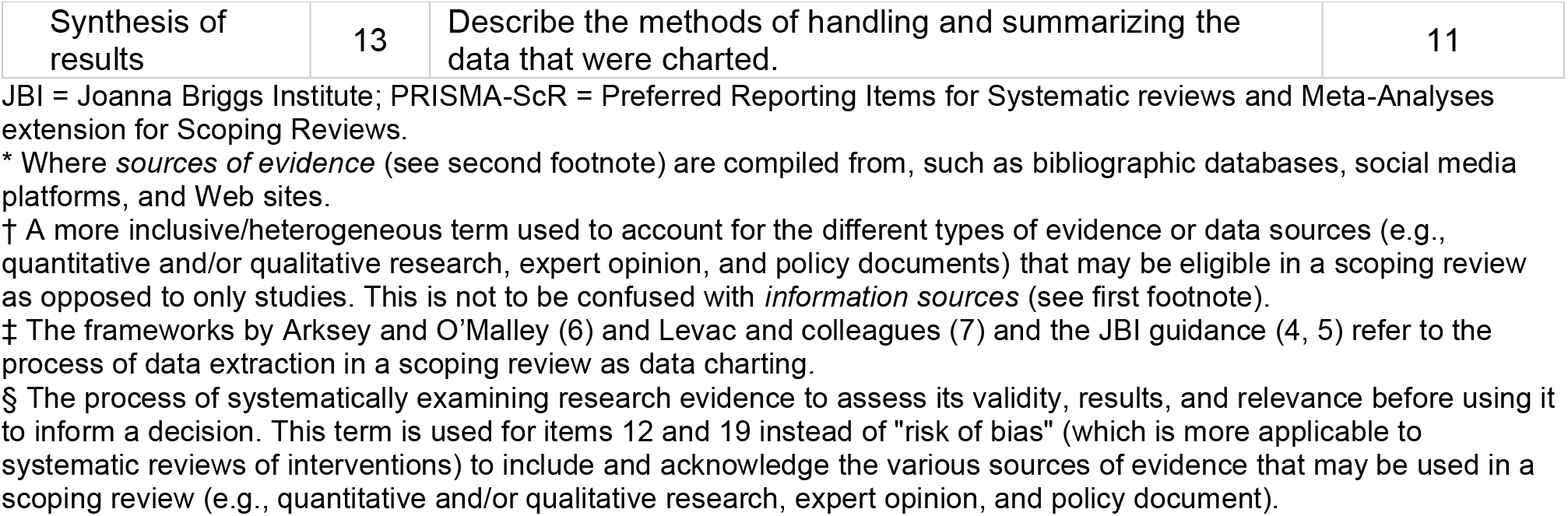

## SUPPLEMENTARY FILE C

**Search strategy**

(“Depressive Disorder”[MeSH Terms] OR “depression”[MeSH Terms]) AND (“Mental Health Services”[MeSH Terms:noexp] OR “Health Care Sector”[MeSH Terms] OR “Organization and Administration”[MeSH Terms] OR “Patient Care Management”[MeSH Terms] OR “Community Health Services”[MeSH Terms] OR “Continuity of Patient Care”[MeSH Terms] OR “General Practice”[MeSH Terms] OR (“mental health service*”[Text Word] OR “primary care”[Text Word] OR “primary health care”[Text Word] OR “secondary care”[Text Word] OR “secondary health care”[Text Word] OR “General Practice”[Text Word] OR “patient discharge”[Text Word] OR “patient transfer*”[Text Word] OR “transitional care”[Text Word] OR “after care”[Text Word] OR “patient care continuity”[Text Word] OR “transition*”[Text Word] OR “health care”[Text Word] OR “Organization”[Text Word])) AND (“Behavioral Disciplines and Activities”[MeSH Terms] OR “Mental Processes”[MeSH Terms] OR “Transtheoretical Model”[MeSH Terms] OR “Therapeutics”[MeSH Terms] OR (“intervention*”[Text Word] OR “method*”[Text Word] OR “model*”[Text Word] OR “procedure*”[Text Word] OR “process*”[Text Word] OR “treatment*”[Text Word] OR “therapy*”[Text Word])) AND (“Mental Health Recovery”[MeSH Terms] OR “Recovery of Function”[MeSH Terms] OR “Return to Work”[MeSH Terms] OR “Return to School”[MeSH Terms] OR “recover*”[Text Word])

## REFERENCES

1. Kessler, R.C., et al., The epidemiology of major depressive disorder: results from the National Comorbidity Survey Replication (NCS-R). Jama, 2003. 289(23): p. 3095–105.

2. Malhi, G.S. and J.J. Mann, Depression. Lancet, 2018. 392(10161): p. 2299–2312.

3. Evaluation, I.o.H.M.a. Global Health Data Exchange (GHDx). [cited 2022 09-05]; Available from: http://ghdx.healthdata.org/gbd-results-tool?params=gbd-api-2019-permalink/d780dffbe8a381b25e1416884959e88b.

4. Executive, B., Global burden of mental disorders and the need for a comprehensive, coordinated response from health and social workers at the country level: report by the Secretariat. 2012, World Health Organization: Geneva.

5. Šprah, L., et al., Psychiatric readmissions and their association with physical comorbidity: a systematic literature review. BMC Psychiatry, 2017. 17(1): p. 2.

6. Plana-Ripoll, O., et al., A comprehensive analysis of mortality-related health metrics associated with mental disorders: a nationwide, register-based cohort study. Lancet, 2019. 394(10211): p. 1827–1835.

7. Steffen, A., et al., Mental and somatic comorbidity of depression: a comprehensive cross-sectional analysis of 202 diagnosis groups using German nationwide ambulatory claims data. BMC Psychiatry, 2020. 20(1): p. 142.

8. Kessler, R.C., K.R. Merikangas, and P.S. Wang, Prevalence, comorbidity, and service utilization for mood disorders in the United States at the beginning of the twenty-first century. Annu Rev Clin Psychol, 2007. 3: p. 137–58.

9. Kaufman, J. and D. Charney, Comorbidity of mood and anxiety disorders. Depress Anxiety, 2000. 12 Suppl 1: p. 69–76.

10. Laursen, T.M., et al., Mortality and life expectancy in persons with severe unipolar depression. J Affect Disord, 2016. 193: p. 203–7.

11. Bock, C., et al., The influence of comorbid personality disorder and neuroticism on treatment outcome in first episode depression. Psychopathology, 2010. 43(3): p. 197–204.

12. Katon, W., E.H. Lin, and K. Kroenke, The association of depression and anxiety with medical symptom burden in patients with chronic medical illness. Gen Hosp Psychiatry, 2007. 29(2): p. 147–55.

13. Carney, R.M. and K.E. Freedland, Depression and coronary heart disease. Nat Rev Cardiol, 2017. 14(3): p. 145–155.

14. Sartorius, N., Depression and diabetes. Dialogues Clin Neurosci, 2018. 20(1): p. 47–52.

15. Kessler, R.C., The costs of depression. Psychiatr Clin North Am, 2012. 35(1): p. 1–14.

16. Saris, I.M.J., et al., Default Mode Network Connectivity and Social Dysfunction in Major Depressive Disorder. Sci Rep, 2020. 10(1): p. 194.

17. Sarris, J., et al., Lifestyle medicine for depression. BMC Psychiatry, 2014. 14: p. 107.

18. Herrman, H., et al., Time for united action on depression: a Lancet-World Psychiatric Association Commission. The Lancet, 2022. 399(10328): p. 957–1022.

19. Liu, Q., et al., Changes in the global burden of depression from 1990 to 2017: Findings from the Global Burden of Disease study. J Psychiatr Res, 2020. 126: p. 134–140.

20. Sheehan, D.V., et al., The Mini-International Neuropsychiatric Interview (M.I.N.I.): the development and validation of a structured diagnostic psychiatric interview for DSM-IV and ICD-10. J Clin Psychiatry, 1998. 59 Suppl 20: p. 22–33;quiz 34-57.

21. England MJ S.L., Depression in Parents, Parenting, and Children: Opportunities to Improve Identification, Treatment, and Prevention. 2009, Washington (DC): National Academies Press (US): National Research Council (US) and Institute of Medicine (US) Committee on Depression, Parenting Practices, and the Healthy Development of Children.

22. Organization, W.H., Depression and Other Common Mental Disorders: Global Health Estimates. 2017:Geneva. p. 24.

23. Weihs, K. and J.M. Wert, A primary care focus on the treatment of patients with major depressive disorder. Am J Med Sci, 2011. 342(4): p. 324–30.

24. Fleury, M.J., et al., General practitioners’ management of mental disorders: a rewarding practice with considerable obstacles. BMC Fam Pract, 2012. 13: p. 19.

25. Murray, R.M., et al., Essential psychiatry. 2008.

26. Cox, G.R., et al., Interventions for preventing relapse and recurrence of a depressive disorder in children and adolescents. Cochrane Database Syst Rev, 2012. 11(11): p. Cd007504.

27. Machmutow, K., et al., Comparative effectiveness of continuation and maintenance treatments for persistent depressive disorder in adults. Cochrane Database Syst Rev, 2019. 5(5): p. Cd012855.

28. Licht, R.W., Is it possible to evaluate true prophylactic efficacy of antidepressants in severely ill patients with recurrent depression? Lessons from a placebo-controlled trial. The fifth trial of the Danish University Antidepressant Group (DUAG-5). J Affect Disord, 2013. 148(2-3): p. 286–90.

29. Hvenegaard, M., et al., Group rumination-focused cognitive-behavioural therapy (CBT) v. group CBT for depression: phase II trial. Psychol Med, 2020. 50(1): p. 11–19.

30. Martiny, K., et al., Maintained superiority of chronotherapeutics vs. exercise in a 20-week randomized follow-up trial in major depression. Acta Psychiatr Scand, 2015. 131(6): p. 446–57.

31. Burcusa, S.L. and W.G. Iacono, Risk for recurrence in depression. Clin Psychol Rev, 2007. 27(8): p. 959–85.

32. Baldessarini, R.J., et al., Illness risk following rapid versus gradual discontinuation of antidepressants. Am J Psychiatry, 2010. 167(8): p. 934–41.

33. Sim, K., et al., Prevention of Relapse and Recurrence in Adults with Major Depressive Disorder: Systematic Review and Meta-Analyses of Controlled Trials. Int J Neuropsychopharmacol, 2015. 19(2).

34. Kessing, L.V. and P.K. Andersen, The effect of episodes on recurrence in affective disorder: a case register study. J Affect Disord, 1999. 53(3): p. 225–31.

35. Kupfer, D.J., Long-term treatment of depression. J Clin Psychiatry, 1991. 52 Suppl: p. 28–34.

36. Tønning, M.L., et al., The effect of smartphone-based monitoring and treatment on the rate and duration of psychiatric readmission in patients with unipolar depressive disorder: The RADMIS randomized controlled trial. J Affect Disord, 2021. 282: p. 354–363.

37. Buckman, J.E.J., et al., Risk factors for relapse and recurrence of depression in adults and how they operate: A four-phase systematic review and meta-synthesis. Clin Psychol Rev, 2018. 64: p. 13–38.

38. Moriarty, A.S., et al., Predicting and preventing relapse of depression in primary care. Br J Gen Pract, 2020. 70(691): p. 54–55.

39. Gaynes, B.N., W.C. Jackson, and K.D. Rorie, Major Depressive Disorder in the Primary Care Setting: Strategies to Achieve Remission and Recovery. J Fam Pract, 2015. 64(9): p. S4–s15.

40. Rait, G., et al., Recent trends in the incidence of recorded depression in primary care. Br J Psychiatry, 2009. 195(6): p. 520–4.

41. Qin, P. and M. Nordentoft, Suicide risk in relation to psychiatric hospitalization: evidence based on longitudinal registers. Arch Gen Psychiatry, 2005. 62(4): p. 427–32.

42. Hansen, H.V., et al., The effects of centralised and specialised intervention in the early course of severe unipolar depressive disorder: a randomised clinical trial. PLoS One, 2012. 7(3): p. e32950.

43. Bosman, R.C., et al., Long-term antidepressant use: a qualitative study on perspectives of patients and GPs in primary care. Br J Gen Pract, 2016. 66(651): p. e708–19.

44. Anthony, W.A., Recovery from mental illness: The guiding vision of the mental health service system in the 1990s. Psychosocial Rehabilitation Journal, 1993. 16(4): p. 11–23.

45. Jørgensen, K., et al., Recovery-oriented intersectoral care between mental health hospitals and community mental health services: An integrative review. Int J Soc Psychiatry, 2020: p. 20764020966634.

46. Saxena, S., M.K. Funk, and D. Chisholm, Comprehensive mental health action plan 2013-2020. East Mediterr Health J, 2015. 21(7): p. 461–3.

47. Rossi, A., et al., The complex relationship between self-reported ‘personal recovery’ and clinical recovery in schizophrenia. Schizophr Res, 2018. 192: p. 108–112.

48. Slade, M., et al., REFOCUS Trial: protocol for a cluster randomised controlled trial of a pro-recovery intervention within community based mental health teams. BMC Psychiatry, 2011. 11: p. 185.

49. Turton, P., et al., One size fits all: or horses for courses? Recovery-based care in specialist mental health services. Soc Psychiatry Psychiatr Epidemiol, 2011. 46(2): p. 127–36.

50. Leamy, M., et al., Conceptual framework for personal recovery in mental health: systematic review and narrative synthesis. Br J Psychiatry, 2011. 199(6): p. 445–52.

51. Bird, V., et al., Fit for purpose? Validation of a conceptual framework for personal recovery with current mental health consumers. Aust N Z J Psychiatry, 2014. 48(7): p. 644–53.

52. Stickley, T., N. Wright, and M. Slade, The art of recovery: outcomes from participatory arts activities for people using mental health services. J Ment Health, 2018. 27(4): p. 367–373.

53. Williams, J., et al., Development and evaluation of the INSPIRE measure of staff support for personal recovery. Soc Psychiatry Psychiatr Epidemiol, 2015. 50(5): p. 777–86.

54. Fava, G.A., C. Ruini, and C. Belaise, The concept of recovery in major depression. Psychol Med, 2007. 37(3): p. 307–17.

55. Anthony, W.A., A recovery-oriented service system: Setting some system level standards. Psychiatric Rehabilitation Journal, 2000. 24(2): p. 159–168.

56. Davidson, L., et al., The top ten concerns about recovery encountered in mental health system transformation. Psychiatr Serv, 2006. 57(5): p. 640–5.

57. Deegan, P.E., Recovery: The lived experience of rehabilitation. Psychosocial Rehabilitation Journal, 1988. 11(4): p. 11–19.

58. Davidsen, A.S., et al., Experiences of barriers to trans-sectoral treatment of patients with severe mental illness. A qualitative study. Int J Ment Health Syst, 2020. 14(1): p. 87.

59. Gili, M., et al., Interventions for preventing relapse or recurrence of depression in primary health care settings: A systematic review. Prev Med, 2015. 76 Suppl: p. S16–21.

60. Katon, W., et al., A randomized trial of relapse prevention of depression in primary care. Arch Gen Psychiatry, 2001. 58(3): p. 241–7.

61. Strand, M., D. Gammon, and C.M. Ruland, Transitions from biomedical to recovery-oriented practices in mental health: a scoping review to explore the role of Internet-based interventions. BMC Health Serv Res, 2017. 17(1): p. 257.

62. Winsper, C., et al., How do recovery-oriented interventions contribute to personal mental health recovery? A systematic review and logic model. Clin Psychol Rev, 2020. 76: p. 101815.

63. Blasi, P.R., et al., Transitioning patients from outpatient mental health services to primary care: A rapid literature review. Implementation Research and Practice, 2021. 2: p. 26334895211041294.

64. Gunn, J., et al., A systematic review of complex system interventions designed to increase recovery from depression in primary care. BMC Health Serv Res, 2006. 6: p. 88.

65. Sheehan, D.V., et al., Restoring function in major depressive disorder: A systematic review. J Affect Disord, 2017. 215: p. 299–313.

66. Richardson, K. and M. Barkham, Recovery from depression: a systematic review of perceptions and associated factors. J Ment Health, 2020. 29(1): p. 103–115.

67. Smith, S.M., et al., Shared care across the interface between primary and specialty care in management of long term conditions. Cochrane Database Syst Rev, 2017. 2(2): p. Cd004910.

68. O’Cathain, A., et al., Guidance on how to develop complex interventions to improve health and healthcare. BMJ Open, 2019. 9(8): p. e029954.

69. Craig, P., et al., Developing and evaluating complex interventions: the new Medical Research Council guidance. Bmj, 2008. 337: p. a1655.

70. Skivington, K., et al., A new framework for developing and evaluating complex interventions: update of Medical Research Council guidance. BMJ, 2021. 374: p. 2061.

71. Arksey, H. and L. O’Malley, Scoping studies: towards a methodological framework. International Journal of Social Research Methodology, 2005. 8(1): p. 19–32.

72. Levac, D., H. Colquhoun, and K.K. O’Brien, Scoping studies: advancing the methodology. Implement Sci, 2010. 5: p. 69.

73. Peters, M.D., et al., Guidance for conducting systematic scoping reviews. Int J Evid Based Healthc, 2015. 13(3): p. 141–6.

74. Peters, M.D.J., et al., Updated methodological guidance for the conduct of scoping reviews. JBI Evid Synth, 2020. 18(10): p. 2119–2126.

75. Grimshaw, J. A guide to knowledge synthesis: a knowledge synthesis chapter. Available from: https://cihr-irsc.gc.ca/e/41382.html.

76. Peters, M., et al., Joanna Briggs Institute reviewer’s manual. The Joanna Briggs Institute, 2017.

77. Peters, M.D.J., et al., Best practice guidance and reporting items for the development of scoping review protocols. JBI Evid Synth, 2022.

78. Tricco, A.C., et al., PRISMA Extension for Scoping Reviews (PRISMA-ScR): Checklist and Explanation. Ann Intern Med, 2018. 169(7): p. 467–473.

79. Munthe-Kaas, H., et al., The TRANSFER Approach for assessing the transferability of systematic review findings. BMC Med Res Methodol, 2020. 20(1): p. 11.

80. Page, M.J., et al., The PRISMA 2020 statement: an updated guideline for reporting systematic reviews. Bmj, 2021. 372: p. 71.

81. Lenzen, S.A., et al., Disentangling self-management goal setting and action planning: A scoping review. PLoS One, 2017. 12(11): p. e0188822.

82. Concannon, T.W., et al., A new taxonomy for stakeholder engagement in patient-centered outcomes research. J Gen Intern Med, 2012. 27(8): p. 985–91.

